# The direct economic impact of surgical non-response in orthopaedic hip, knee, and spine surgery for osteoarthritis: a cost-utility analysis

**DOI:** 10.64898/2026.06.18.26355936

**Authors:** Y. Raja Rampersaud, Anthony V. Perruccio, Emily Collett, Kala Sundararajan, Jin Tong Du, Luis Montoya, J. Denise Power, Mayilee Canizares, Mohit Kapoor, J. Roderick Davey, Rajiv Gandhi, Stephen Lewis, Khalid A. Syed, Christian J. Veillette, Peter C. Coyte, Nizar N. Mahomed

**Author notes:** Corresponding Author: (RR).

## Abstract

**Background:** Annually, nearly 2 million hip, knee, and spinal inpatient surgeries are performed in Canada and the US for osteoarthritis (OA), costing over $37 billion in hospital expenditures. However, 15-30% of patients experience limited or no improvement, resulting in poor value for money. This study evaluated the one-year cost-utility of joint and spine procedures for OA by comparing non-responders to responders, considering various responder definitions.

**Methods:** Individual micro-costing data were collected for 1,175 elective hip, knee, and spine patients enrolled in the Longitudinal Evaluation in the Arthritis Program – Osteoarthritis (LEAP-OA) between 2014 and 2018. Quality-adjusted life years (QALYs) were derived using the SF-6D utility index. One-year incremental cost-utility ratios (ICURs) were calculated from the hospital perspective.

**Results:** Responder rates varied by definition, ranging from 78%-94% for hip replacements, 64%-90% for knee replacements, 60%-64% for spine fusions, and 50%-68% for spine decompressions. Corresponding ICURs were: $45,956-$51,773/QALY for responders versus $108,593-$485,762/QALY for non-responders for hip replacements; $54,831-$71,151/QALY for responders versus $200,486-$1,203,596/QALY for non-responders for knee replacements; $65,980-$74,422/QALY for responders versus $262,039-$729,686/QALY for non-responders for spine fusions; and $29,947-$42,168/QALY for responders versus $63,195-$662,586/QALY for non-responders for spine decompressions.

**Conclusions:** While surgical response rates were highly dependent on the responder definition, ICURs for non-responders were significantly higher than those for responders across all definitions. Beyond the negative impact on patients, there is a compelling economic argument for investment in improved pre-operative identification of patients at risk of surgical non-response. Such efforts could enable more personalized, value-based care pathways and reduce the provision of low-value surgical interventions.

## Introduction

Osteoarthritis (OA) is a leading cause of years lived with disability globally [1]. The prevalence of OA is projected to rise significantly in the coming decades, driven by both the aging population and non-age-related factors [2–6]. For instance, the number of individuals living with OA is expected to increase by 50% in Canada from 2015 to 2040 [4], and by 47% in the US from 2012 to 2040 [7]. This escalating prevalence poses substantial challenges to healthcare systems already grappling with constrained resources.

Surgical interventions for end-stage OA are common, and their growing volumes drive rising healthcare expenditures [8–13]. According to the US Healthcare Cost and Utilization Project (HCUP), hip replacements, knee replacements, and spine fusions were the three most costly procedures in 2018 [14]. From 2008 to 2018, inpatient knee replacements increased by 8.4% to over 705,000 procedures, exceeding $11.8 billion; hip replacements increased by 41.0% to over 591,000 procedures, exceeding $10.4 billion. Spinal fusion, commonly performed for spinal stenosis as a result of OA, increased 5.4%, reaching over 418,000 procedures costing over $14.1 billion [15]. Similarly, in Canada, hip and knee replacements are the fastest-growing surgeries [11,16]. In 2018, 75,345 knee replacements represented a 22.5% increase over five years, while 62,016 hip replacements marked a 20.1% rise, collectively costing more than $1.4 billion annually [17].

While OA surgeries are generally considered cost-effective in improving pain and function [18–21], 15-30% of patients experience limited or no improvement, termed "non-responders" [22–26]. The concept of responder status in OA surgery is not universally defined [27]. Various criteria, such as achieving a minimally clinically important difference (MCID) [28–31], overall satisfaction [32], or attaining a patient acceptable symptom state (PASS) [33], have been used in the literature. Irrespective of the definition, non-response rates average ∼20%, with hip replacement generally outperforming knee replacement and spinal surgeries [34–36]. The personal and economic implications of surgical non-response are substantial. Non-responders not only experience continued pain and functional limitations, but also represent a significant financial burden on the healthcare system due to suboptimal outcomes despite the initial surgical investment (i.e. low-value to both payer and patient) [32,37].

The objective of this study was to evaluate the one-year cost-utility of hip, knee, and spine surgeries for OA by comparing non-responders to responders across different definitions. We highlight the health economic consequences of surgical non-response, providing a foundation for future efforts to improve patient selection and outcome prediction in OA surgical care.

## Methods

### Study Design and Patient Population

A retrospective cost-utility analysis was performed using data from the Longitudinal Evaluation in the Arthritis Program – Osteoarthritis (LEAP-OA), a prospective cohort study (REB#16-5759) conducted at the University Health Network (UHN). Elective hip replacement, knee replacement, 1-3 level spine fusion with or without decompression, and 1-3 level spine decompression alone for primary OA between November 18, 2013 and March 12, 2018 were included. Data were accessed for analysis on June 8, 2025.

### Data Collection

#### Outcome measures

Patients completed questionnaires within 30 days pre-operatively and at one year post-operatively. Patient-reported outcome measures (PROMs) included the Western Ontario and McMaster Universities (WOMAC) Osteoarthritis Index pain (range 0-20) and function (range 0-68) subscales for hip and knee OA; the Oswestry Disability Index (ODI; range 0-100) for spine conditions, with higher scores indicating worse pain or disability; and the Numeric Pain Rating Scale (NPRS; range 0-10), with higher scores indicating worse pain. Generic measures included the SF-12 Physical Component Summary (PCS; range 0-100), where higher scores indicated better health. Patients also answered the PASS question and rated their overall satisfaction with surgery.

Six PROM-based responder definitions were considered at one-year post-operation (Table 1).

**Table 1.** Summary of joint-specific and generic responder definitions at one-year post-operation.

| Category | Measure | Definition |  |
| --- | --- | --- | --- |
| Joint-specific | Percentage Improvement | Hip | ≥30% improvement in WOMAC pain or function scores |
|  |  | Knee |  |
|  |  | Spine | ≥30% improvement in ODI scores |
|  | MCID | Hip | ≥29.26-point improvement in WOMAC pain and ≥26.54-point improvement in WOMAC function [30] |
|  |  | Knee | ≥22.87-point improvement in WOMAC pain and ≥19.01-point improvement in WOMAC function [29] |
|  |  | Spine | ≥12.8-point improvement in ODI [31] |
|  | PASS | Answering “Yes” to “*Taking into account how you are now, can you cope with the current state of your [joint] and do everything you need to do in your daily life?*” |  |
|  | Pain Reduction | ≥3-point reduction in the NPRS [38,39], based on the item “*Please rate the intensity of the average pain in your [joint] during the past week.*” scored on a 0-10 numeric rating scale (0 = no pain, 10 = worst pain imaginable). |  |
| Generic | Physical Health Improvement | ≥4.6-point improvement in SF-12 PCS [40] |  |
|  | Patient Satisfaction | Reporting “Satisfied” (very or somewhat satisfied) versus “not satisfied” (somewhat or very dissatisfied) with surgery |  |

Abbreviations: WOMAC = Western Ontario and McMaster Universities Osteoarthritis Index;
ODI = Oswestry Disability Index; MCID = Minimal Clinically Important Difference; PASS =
Patient Acceptable Symptom State; NPRS = Numeric Pain Rating Scale; SF-12 PCS = Short
Form-12 Physical Component Summary.

#### Cost data

Hospital micro-case cost data ($CAD) were obtained from the UHN Case Costing & Activity Reporting Department as a part of the Ontario Case Costing Initiative (OCCI) [41]. Total hospital costs were captured for the index episode of care (from admission to discharge) for each patient and reflect the costs incurred in the year the surgery was performed. Direct costs encompassed labour, operating and recovery room hours, surgical implants, disposables, ward nursing and allied healthcare, inpatient diagnostic imaging, lab, and pharmacy charges. Indirect costs, such as administrative and facilities costs, were also included.

### Cost-Utility Analysis

One-year incremental cost-utility ratios (ICURs) were calculated from the hospital’s perspective, representing the total hospital costs ($CAD) per quality-adjusted life year (QALY) gained following surgery. QALYs were derived using the SF-6D utility index (range 0.29–1.00; 0.29 = worst health, 1.00 = full health) based on the SF-12 survey. As all patients in the cohort underwent surgery, a conservative assumption was made that, in the absence of surgery, patients would have incurred no additional hospital costs and experienced no change in health-related quality of life over the one-year time horizon. This reflects typical preoperative clinical stability during prolonged wait times from referral for surgical assessment to surgery [42–45], and provides a reference for interpreting cost-utility outcomes. ICUR was calculated as 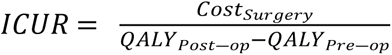.

### Statistical Analysis

Descriptive statistics summarized the hip, knee and spine cohorts. Responder rates were calculated by procedure and definition. Mean incremental costs, QALYs gained, and ICURs were compared between responders and non-responders by procedure.

## Results

A total of 1,175 patients were included in the baseline cohort (501 hip replacements, 541 knee replacements, 45 spine fusions, and 88 spine decompressions). The mean age was 65.6 years, and 54.5% of participants were female. Table 2 summarizes the baseline sociodemographic profile and the baseline and one-year PROMs by procedure subgroups.

**Table 2.**
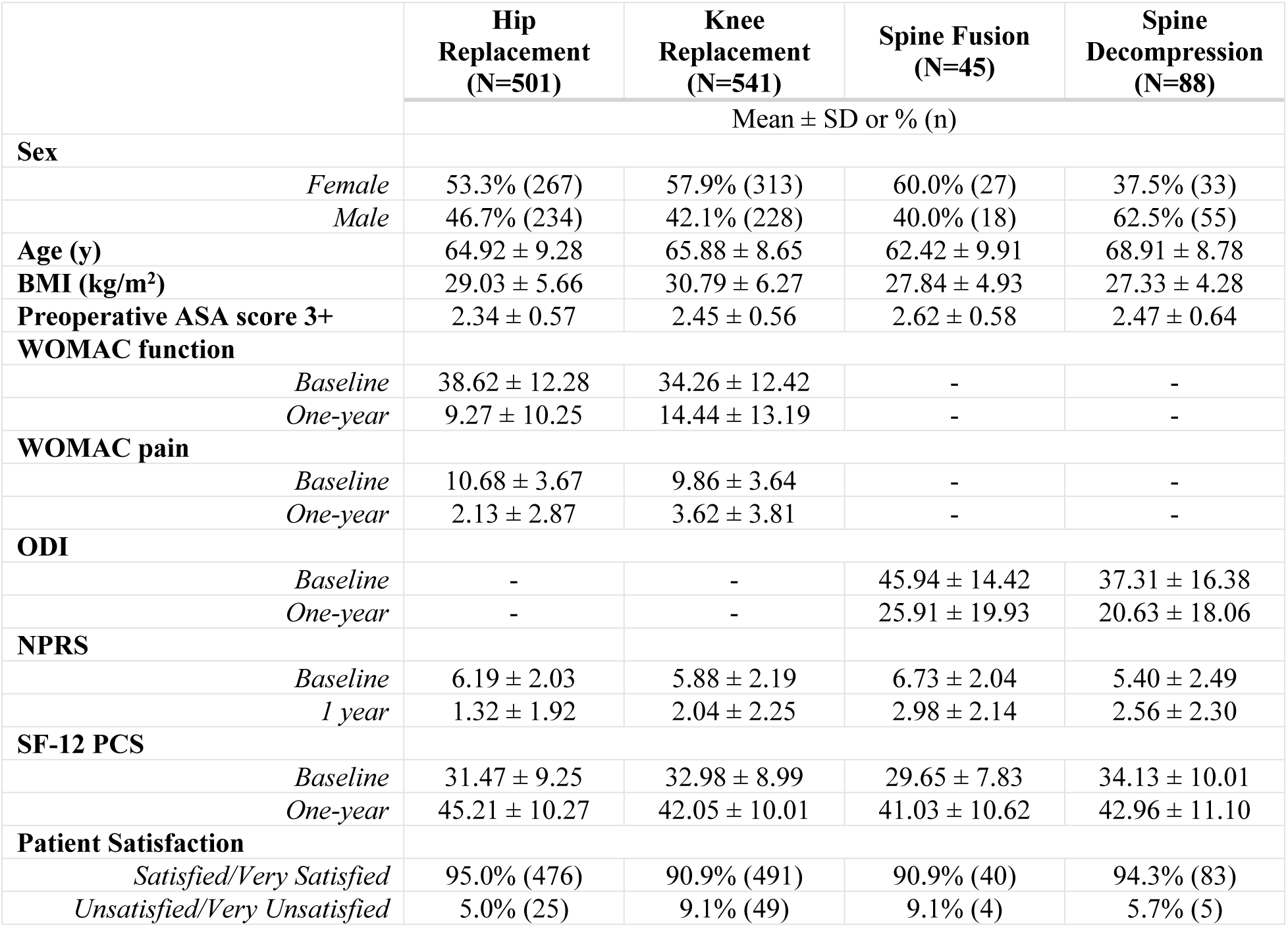
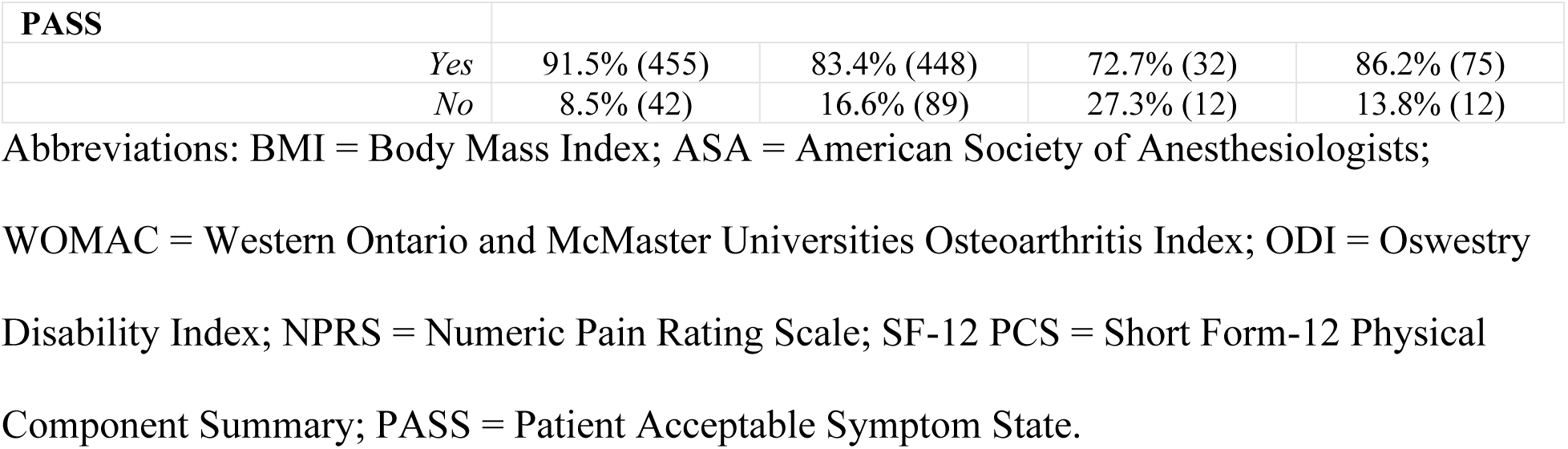
Baseline sociodemographic characteristics and patient-reported outcome measures at baseline and one-year post-operation.

|  |  | **Hip**<br>**Replacement**<br>**(N=501)** | **Knee**<br>**Replacement**<br>**(N=541)** | **Spine Fusion**<br>**(N=45)** | **Spine**<br>**Decompression**<br>**(N=88)** |
| --- | --- | --- | --- | --- | --- |
| | | Mean ± SD or % (n) | | | |
| **Sex** |  |  |  |  |  |
|  | *Female* | 53.3% (267) | 57.9% (313) | 60.0% (27) | 37.5% (33) |
|  | *Male* | 46.7% (234) | 42.1% (228) | 40.0% (18) | 62.5% (55) |
| **Age (y)** | | 64.92 ± 9.28 | 65.88 ± 8.65 | 62.42 ± 9.91 | 68.91 ± 8.78 |
| **BMI (kg/m<sup>2</sup>)** | | 29.03 ± 5.66 | 30.79 ± 6.27 | 27.84 ± 4.93 | 27.33 ± 4.28 |
| **Preoperative ASA score 3+** | | 2.34 ± 0.57 | 2.45 ± 0.56 | 2.62 ± 0.58 | 2.47 ± 0.64 |
| **WOMAC function** |  |  |  |  |  |
| | *Baseline* | 38.62 ± 12.28 | 34.26 ± 12.42 | - | - |
| | *One-year* | 9.27 ± 10.25 | 14.44 ± 13.19 | - | - |
| **WOMAC pain** |  |  |  |  |  |
| | *Baseline* | 10.68 ± 3.67 | 9.86 ± 3.64 | - | - |
| | *One-year* | 2.13 ± 2.87 | 3.62 ± 3.81 | - | - |
| **ODI** |  |  |  |  |  |
| | *Baseline* | - | - | 45.94 ± 14.42 | 37.31 ± 16.38 |
| | *One-year* | - | - | 25.91 ± 19.93 | 20.63 ± 18.06 |
| **NPRS** |  |  |  |  |  |
| | *Baseline* | 6.19 ± 2.03 | 5.88 ± 2.19 | 6.73 ± 2.04 | 5.40 ± 2.49 |
| | *1 year* | 1.32 ± 1.92 | 2.04 ± 2.25 | 2.98 ± 2.14 | 2.56 ± 2.30 |
| **SF-12 PCS** |  |  |  |  |  |
| | *Baseline* | 31.47 ± 9.25 | 32.98 ± 8.99 | 29.65 ± 7.83 | 34.13 ± 10.01 |
| | *One-year* | 45.21 ± 10.27 | 42.05 ± 10.01 | 41.03 ± 10.62 | 42.96 ± 11.10 |
| **Patient Satisfaction** |  |  |  |  |  |
|  | *Satisfied/Very Satisfied* | 95.0% (476) | 90.9% (491) | 90.9% (40) | 94.3% (83) |
|  | *Unsatisfied/Very Unsatisfied* | 5.0% (25) | 9.1% (49) | 9.1% (4) | 5.7% (5) |

| PASS |  |  |  |  |  |
| --- | --- | --- | --- | --- | --- |
|  | *Yes* | 91.5% (455) | 83.4% (448) | 72.7% (32) | 86.2% (75) |
|  | *No* | 8.5% (42) | 16.6% (89) | 27.3% (12) | 13.8% (12) |

Abbreviations: BMI = Body Mass Index; ASA = American Society of Anesthesiologists; WOMAC = Western Ontario and McMaster Universities Osteoarthritis Index; ODI = Oswestry Disability Index; NPRS = Numeric Pain Rating Scale; SF-12 PCS = Short Form-12 Physical Component Summary; PASS = Patient Acceptable Symptom State.

One-year follow-up data were available for 85% of hip, 69% of knee, 83% of spine fusion, and 79% of spine decompression patients. Baseline characteristics were generally similar between those with and without follow-up across all procedures (Table 3). In the spine fusion subgroup, patients lost to follow-up had baseline ODI scores that were 6.19 points lower and PCS scores that were 6.46 points higher compared to those retained. In the spine decompression group, those lost to follow-up had baseline ODI scores 5.11 points higher than those retained.

**Table 3.** Baseline sociodemographic characteristics and patient-reported outcome measures of patients retained versus lost to one-year follow-up.

|  | Hip Replacement |  | Knee Replacement |  | Spine Fusion |  | Spine Decompression |  |
| --- | --- | --- | --- | --- | --- | --- | --- | --- |
|  | Retained<br>(N=501) | No F/U<br>(N=145) | Retained<br>(N=541) | No F/U<br>(N=239) | Retained<br>(N=45) | No F/U<br>(N=9) | Retained<br>(N=88) | No F/U<br>(N=23) |
| | Mean ± SD or % (n) | | | | | | | |
| **Sex** |  |  |  |  |  |  |  |  |
| *Female* | 53.3%<br>(267) | 55.2%<br>(80) | 57.9%<br>(313) | 54.0%<br>(129) | 60.0%<br>(27) | 33.3%<br>(3) | 37.5%<br>(33) | 34.8%<br>(8) |
| *Male* | 46.7%<br>(234) | 44.8%<br>(65) | 42.1%<br>(228) | 46.0%<br>(110) | 40.0%<br>(18) | 66.7%<br>(6) | 62.5%<br>(55) | 65.2%<br>(15) |
| **Age (y)** | 64.92 ±<br>9.28 | 65.54 ±<br>10.29 | 65.88 ±<br>8.65 | 66.62 ±<br>10.11 | 62.42 ±<br>9.91 | 65.44 ±<br>12.14 | 68.91 ±<br>8.78 | 70.78 ±<br>10.93 |
| **BMI (kg/m<sup>2</sup>)** | 29.03 ±<br>5.66 | 28.75 ±<br>6.37 | 30.79 ±<br>6.27 | 31.56 ±<br>6.40 | 27.84 ±<br>4.93 | 27.14 ±<br>3.37 | 27.33 ±<br>4.28 | 26.65 ±<br>3.64 |
| **Preoperative ASA score 3+** | 2.34 ±<br>0.57 | 2.47 ±<br>0.55 | 2.45 ±<br>0.56 | 2.50 ±<br>0.56 | 2.62 ±<br>0.58 | 2.67 ±<br>0.50 | 2.47 ±<br>0.64 | 2.43 ±<br>0.66 |
| **WOMAC Function** | 38.62 ±<br>12.28 | 40.25 ±<br>12.30 | 34.26 ±<br>12.42 | 36.75 ±<br>12.69 | - | - | - | - |
| **WOMAC Pain** | 10.68 ±<br>3.67 | 10.76 ±<br>3.60 | 9.86 ±<br>3.64 | 10.50 ±<br>3.86 | - | - | - | - |
| **ODI** | - | - | - | - | 45.94 ±<br>14.42 | 39.75 ±<br>18.17 | 37.31 ±<br>16.38 | 42.42 ±<br>19.77 |
| **NPRS** | 6.19 ±<br>2.03 | 6.09 ±<br>2.29 | 5.88 ±<br>2.19 | 6.32 ±<br>2.09 | 6.73 ±<br>2.04 | 7.88 ±<br>1.25 | 5.40 ±<br>2.49 | 5.58 ±<br>3.06 |
| **SF-12 PCS** | 31.47 ±<br>9.25 | 31.76 ±<br>9.20 | 32.98 ±<br>8.99 | 32.31 ±<br>8.66 | 29.65 ±<br>7.83 | 36.11 ±<br>13.12 | 34.13 ±<br>10.01 | 32.84 ±<br>9.76 |

Abbreviations: BMI = Body Mass Index; ASA = American Society of Anesthesiologists;
WOMAC = Western Ontario and McMaster Universities Osteoarthritis Index; ODI = Oswestry
Disability Index; NPRS = Numeric Pain Rating Scale; SF-12 PCS = Short Form-12 Physical
Component Summary.

Responder rates varied substantially by definition and procedure (Table 4). Hip replacement had the highest rates (78.2-95.0%), followed by knee replacement (64.0-90.9%), spine fusion (60.0-90.9%), and spine decompression (50.0-94.3%). Across procedures, patient satisfaction yielded the highest responder rates. The lowest rates varied by procedure: MCID for hip and knee replacements, percentage improvement for spine fusion, and NPRS reduction for spine decompression.

**Table 4.**
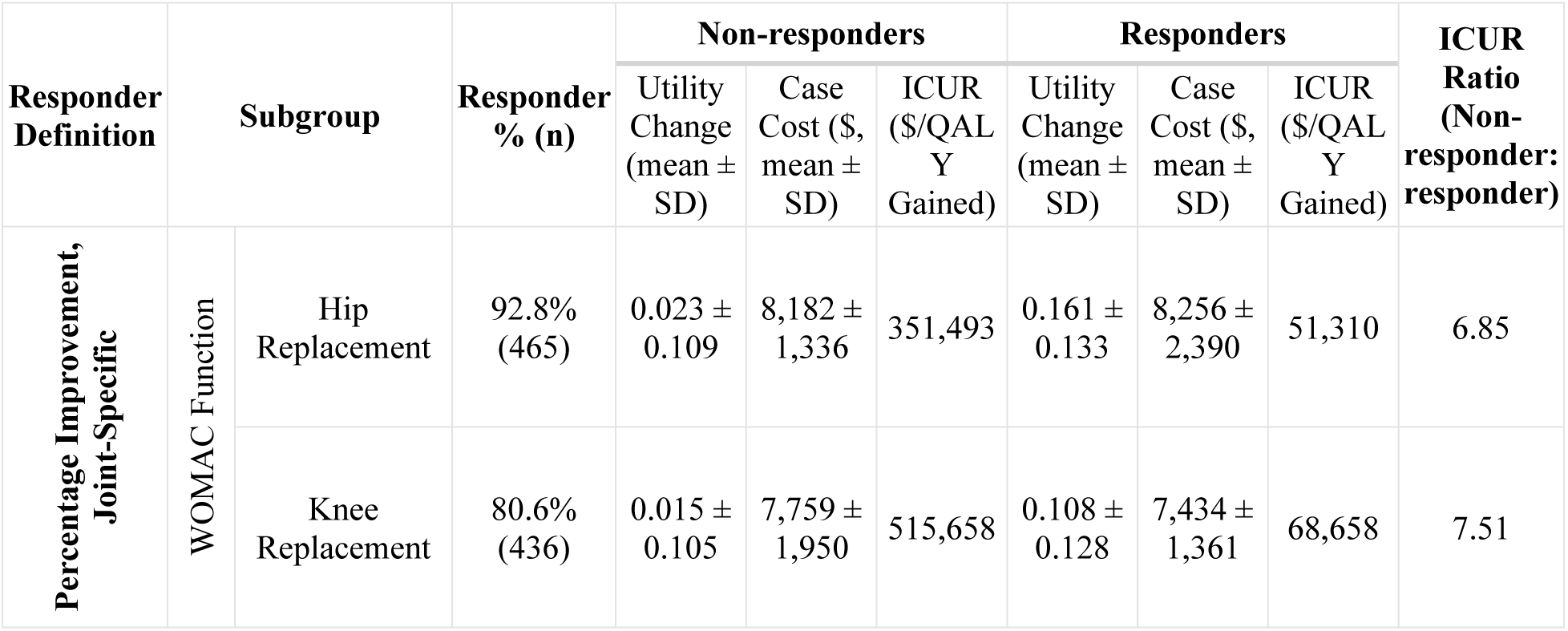
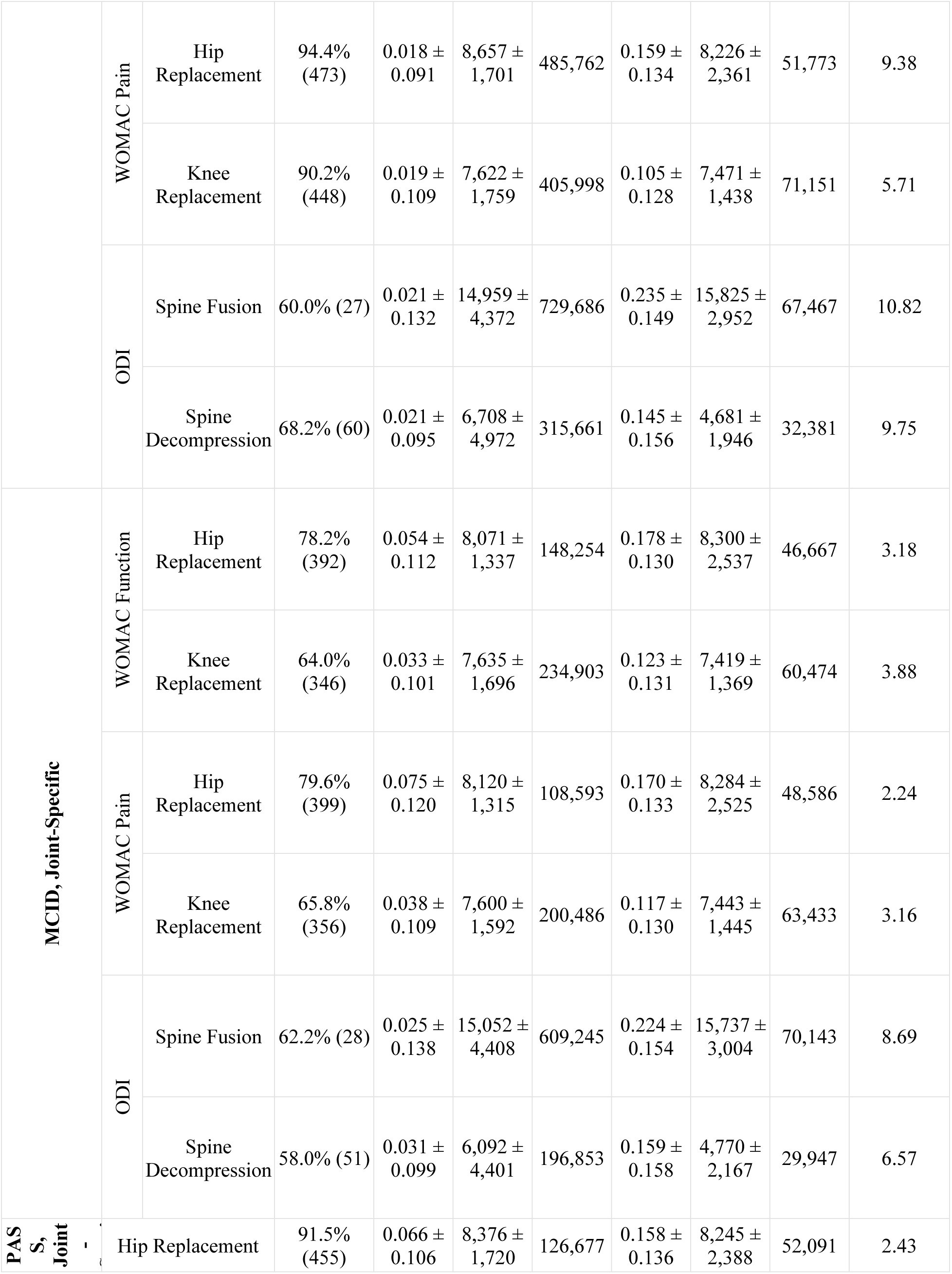
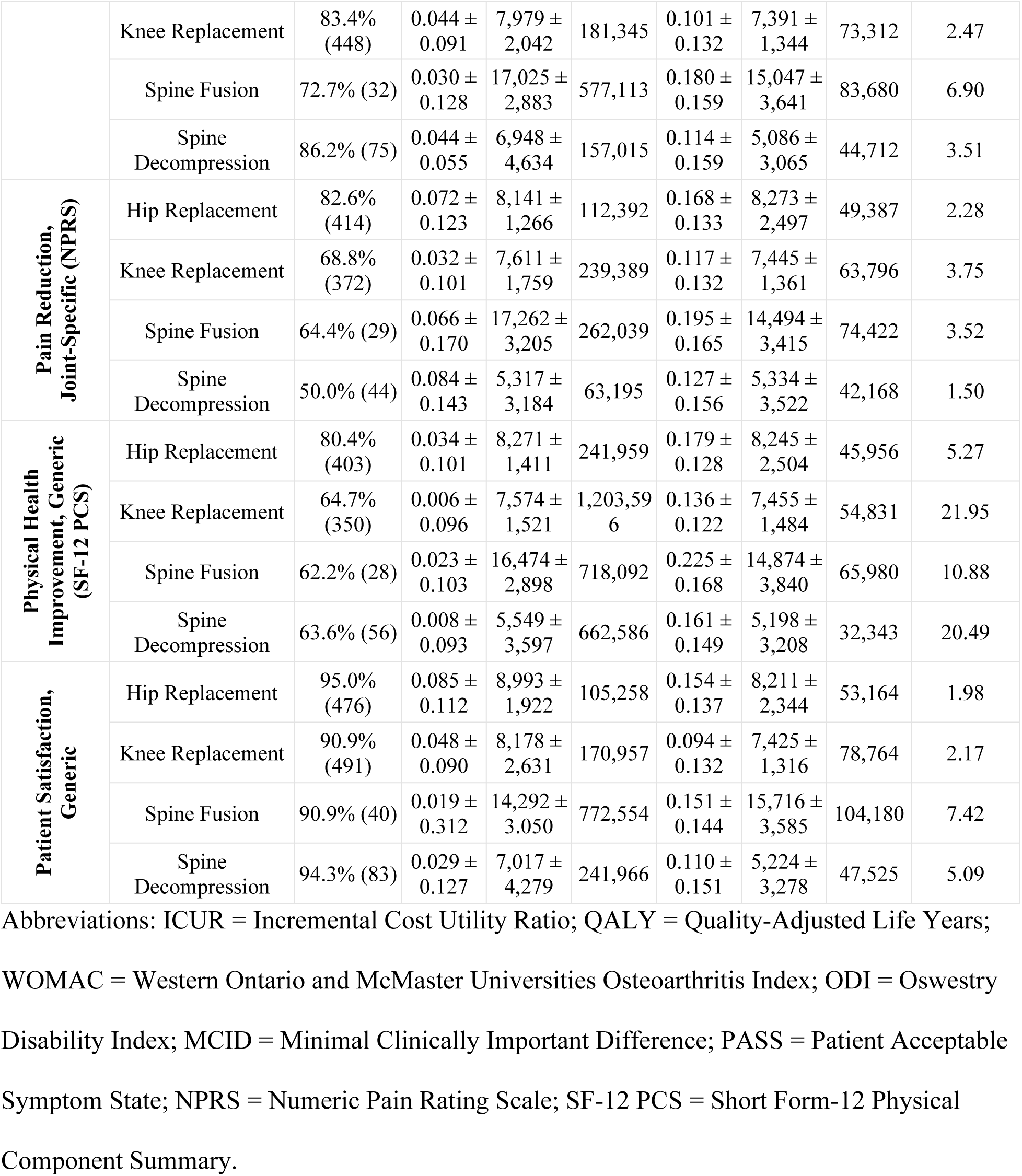
Comparison of one-year change in health utility and hospital costs between responders and non-responders across hip, knee, and spine surgeries for osteoarthritis.

### Cost-Utility Analysis

ICURs varied substantially across procedures (Table 5). Hip replacements had an ICUR of $54,635/QALY, knee replacements $83,135/QALY, spine decompression $50,567/QALY, and spine fusion $103,928/QALY.

**Table 5.** Mean one-year change in health utility and hospital costs across hip, knee, and spine surgeries for osteoarthritis.

| Procedure | N | Utility Change<br>(mean ± SD) | Case Cost<br>(\$, mean ± SD) | ICUR<br>(\$/QALY Gained) |
| --- | --- | --- | --- | --- |
| **Hip** | 501 | 0.151 ± 0.136 | 8,250 ± 2,330 | 54,635 |
| **Knee** | 541 | 0.090 ± 0.129 | 7,497 ± 1,497 | 83,135 |
| **Spine** | 133 | 0.120 ± 0.160 | 8,761 ± 5,902 | 72,960 |
| **Spine Fusion** | 45 | 0.149 ± 0.176 | 15,478 ± 3,566 | 103,928 |
| **Spine Decompression** | 88 | 0.105 ± 0.150 | 5,326 ± 3,338 | 50,567 |
| **All Procedures** | 1,175 | 0.119 ± 0.139 | 7,961 ± 2,732 | 66,622 |

Abbreviations: ICUR = Incremental Cost Utility Ratio; QALY = Quality-Adjusted Life Years.

ICRUs also varied substantially within procedures between responders and non-responders, irrespective of the definition (Table 4). Responders consistently gained more QALYs than non-responders at one-year post-operation, while within-procedure case costs were comparable. ICURs for responders versus non-responders were $45,956-$53,164/QALY versus $105,258-$485,762/QALY for hip replacements, $54,831-$78,764/QALY versus $170,957- $1,203,596/QALY for knee replacements, $65,980-$104,180/QALY versus $262,039- $772,554/QALY for spine fusions, and $29,947-$47,525/QALY versus $63,195- $662,586/QALY for spine decompressions.

## Discussion

Our analysis of four of the most common orthopedic procedures performed in North America [46] highlights significant health economic concerns associated with surgical non-response in hip, knee, and spine surgeries for OA. While surgical response rates were highly definition-dependent, these findings were nevertheless consistent. Despite considerable annual expenditures on these procedures, a notable proportion of patients do not achieve meaningful clinical benefits, resulting in low-value care in approximately one in five surgeries.

Our observed responder rates largely align with the ranges in the existing literature. For hip replacement, our response rates ranged from 78%-95%, similar to those reported by Beswick et al. [23], with long-term pain in 7%-23% of patients using WOMAC pain scores. Knee replacement response rates in our study were 64%-91%, aligning with te Molder et al. [47], who found non-response rates between 4%-42% across 15 definitions, and with Cheng et al. [48], who estimated that 12.6% of patients had persistent pain at one year using NPRS ≥ 4. For spine fusions, our observed responder rates ranged from 60%–91%, which is comparable to reports from Turcotte et al. [49], who found that 69% of patients achieved MCID following lumbar fusion, and Hatakka et al. [50], who reported that 74% of patients achieved a ≥30% ODI improvement. For decompression only, responder rates in our study ranged from 50%–94%, aligning with Bech-Azeddine et al. [51], who found that 59% of patients achieved MCID for ODI, and Sunderland et al. [52], who reported that 24% of patients undergoing primary lumbar decompression were non-responders.

Our study demonstrated that responder rates varied by definition, with MCID and PCS yielding lower rates, while patient satisfaction and PASS yielded the highest rates. Because the responder definitions do not consistently classify the same patients, the composition of the responder and non-responder groups changes across definitions, which likely contributes to the observed differences in ICURs. Nevertheless, the health economic impact remains consistent: OA surgeries that result in non-response represent low-value use of healthcare resources, as they incur comparable hospital costs but achieve minimal or no QALY gain. In contrast, high-value surgery offers benefits, including improved pain and function, at reasonable costs, resulting in a favourable ICUR [53]. In our study, responders fall into the high-value category, demonstrating QALY gains at the same cost as non-responders. While our results do not identify predictors of response, they underscore the need for more active preoperative measures to improve patient selection and enhance the value of surgical care.

Most health economic studies report an average ICUR to reflect the relative value of payer investment for the overall group [54,55]. Our study’s combined average ICUR of $66,622/QALY (Table 5) falls within the commonly accepted thresholds of $50,000-$ 150,000/QALY [56]. However, using this average value masks the critical distinction between responders and non-responders, where the latter’s ICURs significantly exceed conventional thresholds. Given the high volume of these procedures, even low non-response rates result in substantial costs and poor outcomes for a large number of patients. To provide perspective, in 2018, approximately 1.8 million joint and spine procedures were completed in the US, costing over USD $36.4 billion [14], while Canada saw 165,000 procedures [9,17] at a cost of CAD $1.8 billion [17,57]. With an overall conservative non-response rate of 20%, this translates to USD $7.3 billion and CAD $360 million spent annually in the US and Canada, respectively, for low-value outcomes affecting 360,000 Americans and 33,000 Canadians. These estimates only represent inpatient procedures and do not include costs beyond discharge [58], nor include outpatient joint replacement or spinal procedures for OA.

This study adds a critical health economic dimension to the literature on surgical outcomes in OA. While prior research has identified various biopsychosocial factors as prognostic for non-responders, these factors are often studied in isolation, and no single set fully explains post-surgical outcomes [59]. Our findings provide a strong economic justification for intensified investment into developing more comprehensive predictive tools and risk mitigation protocols to optimize outcomes. These priorities are reflected in broader efforts to advance value-based care in orthopedics [60]. Identifying high-risk patients pre-operatively could enable more informed shared decision-making, either by diverting likely non-responders to alternative interventions or by targeting modifiable risk factors pre-operatively to improve the likelihood of response, thus improving healthcare value.

## Limitations

This study has several limitations. First, this study reflects a censored population, including only patients who underwent surgery. As such, findings may not generalize to individuals assessed but not selected for surgery, or those who chose not to proceed. Second, there was missing follow-up data. The most notable differences were observed in the spine fusion group, where patients who were lost to follow-up had lower baseline ODI scores and higher PCS scores. The difference in ODI falls below the established MCID, suggesting it is unlikely to be clinically meaningful. In contrast, the 6.46 point/100 difference in PCS exceeds the commonly accepted MCID threshold of 4.6 points, indicating a potentially meaningful difference in baseline physical health status. Given that greater baseline impairment is typically associated with larger postoperative gains [29–31], these patients may have experienced smaller QALY improvements, making our ICUR estimates for this group conservative. In the decompression subgroup, patients without follow-up reported higher baseline disability, though the difference was below the MCID threshold and therefore unlikely to be clinically meaningful. Third, the analysis was conducted from the hospital’s perspective and excluded costs beyond discharge, such as emergency department visits, readmissions, primary care, rehabilitation, medications, and societal costs, including productivity loss and formal/informal caregiving. Inclusion of these costs would likely further increase the estimated burden of surgical non-response. Fourth, the one-year time horizon, while conservative, already demonstrates a substantial economic gap between responders and non-responders. Given the durability of outcomes in responders and the typical absence of continued QALY gains in non-responders, this discrepancy is likely to widen over time [42,43], reinforcing the need to address surgical non-response to support long-term healthcare sustainability and patient outcomes. Finally, the cohort was drawn from a single academic center within a publicly funded healthcare system, which may limit the generalizability of the findings to other practice settings. Nonetheless, existing literature suggests that PROMs are comparable between academic and community hospitals in the context of joint replacements [61].

## Conclusion

Surgical response rates in high-volume OA procedures vary depending on the clinical criterion used. However, across all definitions and PROMs, non-responders consistently have significantly higher ICURs than responders. These findings highlight the economic importance of further investment in research, development, and implementation of point-of-care tools and clinical pathways to better identify and support OA patients at risk of poor surgical outcomes. By enhancing the ability to match patients with appropriate interventions, such efforts have the potential to reduce the long-term healthcare and economic burden associated with low-value surgical care.

## Data Availability

The minimal data set underlying the findings of this study cannot be shared publicly due to ethical restrictions and patient confidentiality however, the data are available upon reasonable request from the University Health Network Research Ethics Board (contact via the UHN Research Ethics Office) for researchers who meet the criteria for access to confidential data.

## References

1. Vos T, Flaxman AD, Naghavi M, Lozano R, Michaud C, Ezzati M, et al. Years lived with disability (YLDs) for 1160 sequelae of 289 diseases and injuries 1990-2010: a systematic analysis for the Global Burden of Disease Study 2010. Lancet. 2012 Dec 15;380(9859):2163–96. doi:10.1016/S0140-6736(12)61729-2 PubMed PMID: 23245607; PubMed Central PMCID: PMC6350784.

2. Perruccio AV, Power JD, Badley EM. Revisiting arthritis prevalence projections--it’s more than just the aging of the population. J Rheumatol. 2006 Sep;33(9):1856–62. PubMed PMID: 16960946.

3. Canada PHA of. Osteoarthritis in Canada [research] [Internet]. 2020 [cited 2024 Mar 28]. Available from: https://www.canada.ca/en/public-health/services/publications/diseases-conditions/osteoarthritis.html

4. Badley EM, Goulart CM, Millstone DB, Perruccio AV. An Update on Arthritis in Canada — National and Provincial Data Regarding the Past, Present, and Future. The Journal of Rheumatology. 2019 Feb 15. doi:10.3899/jrheum.180147 PubMed PMID: 30770501.

5. Ma W, Chen H, Yuan Q, Chen X, Li H. Global, regional, and national epidemiology of osteoarthritis in working-age individuals: insights from the global burden of disease study 1990–2021. Sci Rep. 2025 Mar 6;15(1):7907. doi:10.1038/s41598-025-91783-6

6. Steinmetz JD, Culbreth GT, Haile LM, Rafferty Q, Lo J, Fukutaki KG, et al. Global, regional, and national burden of osteoarthritis, 1990–2020 and projections to 2050: a systematic analysis for the Global Burden of Disease Study 2021. The Lancet Rheumatology. 2023 Sep 1;5(9):e508–22. doi:10.1016/S2665-9913(23)00163-7 PubMed PMID: 37675071.

7. Hootman JM, Helmick CG, Barbour KE, Theis KA, Boring MA. Updated Projected Prevalence of Self-Reported Doctor-Diagnosed Arthritis and Arthritis-Attributable Activity Limitation Among US Adults, 2015–2040. Arthritis & Rheumatology. 2016;68(7):1582–7. doi:10.1002/art.39692

8. Deyo RA, Mirza SK, Martin BI, Kreuter W, Goodman DC, Jarvik JG. Trends, major medical complications, and charges associated with surgery for lumbar spinal stenosis in older adults. JAMA. 2010 Apr 7;303(13):1259–65. doi:10.1001/jama.2010.338 PubMed PMID: 20371784; PubMed Central PMCID: PMC2885954.

9. Dandurand C, Laghaei PF, Fisher CG, Ailon T, Dvorak M, Kwon BK, et al. Out-of-hours emergent surgery for degenerative spinal disease in Canada: a retrospective cohort study from a national registry. The Lancet Regional Health – Americas. 2024 Aug 1;36. doi:10.1016/j.lana.2024.100816

10. Shichman I, Roof M, Askew N, Nherera L, Rozell JC, Seyler TM, et al. Projections and Epidemiology of Primary Hip and Knee Arthroplasty in Medicare Patients to 2040-2060. JB JS Open Access. 2023 Feb 28;8(1):e22.00112. doi:10.2106/JBJS.OA.22.00112 PubMed PMID: 36864906; PubMed Central PMCID: PMC9974080.

11. Hip and Knee Replacements in Canada: CJRR Annual Report, 2021–2022.

12. Sloan M, Premkumar A, Sheth NP. Projected Volume of Primary Total Joint Arthroplasty in the U.S., 2014 to 2030. J Bone Joint Surg Am. 2018 Sep 5;100(17):1455–60. doi:10.2106/JBJS.17.01617 PubMed PMID: 30180053.

13. Jennison T, MacGregor A, Goldberg A. Hip arthroplasty practice across the Organisation for Economic Co-operation and Development (OECD) over the last decade. annals. 2023 Sep;105(7):645–52. doi:10.1308/rcsann.2022.0101

14. H-CUP. Overview of Operating Room Procedures During Inpatient Stays in U.S. Hospitals, 2018. 2018.

15. U.S. Agency for Healthcare Research and Quality. Healthcare Cost and Utilization Project (HCUP) [Internet]. [cited 2025 Jun 8]. HCUPnet Query: Knee Replacement, Hip Replacement, and Spine Fusion Volumes (2000-2022). Available from: https://datatools.ahrq.gov/hcupnet/

16. Hospital stays in Canada, 2022–2023 | CIHI [Internet]. [cited 2025 Jun 8]. Available from: https://www.cihi.ca/en/hospital-stays-in-canada-2022-2023

17. CJRR. Hip and Knee Replacements in Canada: CJRR Annual Statistics Summary, 2018–2019. 2019.

18. Losina E, Walensky RP, Kessler CL, Emrani PS, Reichmann WM, Wright EA, et al. Cost-effectiveness of Total Knee Arthroplasty in the United States: Patient Risk and Hospital Volume. Archives of Internal Medicine. 2009 Jun 22;169(12):1113–21. doi:10.1001/archinternmed.2009.136

19. Daigle ME, Weinstein AM, Katz JN, Losina E. The cost-effectiveness of total joint arthroplasty: a systematic review of published literature. Best Pract Res Clin Rheumatol. 2012 Oct;26(5):649–58. doi:10.1016/j.berh.2012.07.013 PubMed PMID: 23218429; PubMed Central PMCID: PMC3879923.

20. Agarwal N, To K, Khan W. Cost effectiveness analyses of total hip arthroplasty for hip osteoarthritis: A PRISMA systematic review. International Journal of Clinical Practice. 2021;75(2):e13806. doi:10.1111/ijcp.13806

21. Wilson RA, Gwynne-Jones DP, Sullivan TA, Abbott JH. Total Hip and Knee Arthroplasties Are Highly Cost-Effective Procedures: The Importance of Duration of Follow-Up. The Journal of Arthroplasty. 2021 Jun 1;36(6):1864–1872.e10. doi:10.1016/j.arth.2021.01.038

22. Hawker GA, Bohm E, Dunbar MJ, Faris P, Jones CA, Noseworthy T, et al. Patient appropriateness for total knee arthroplasty and predicted probability of a good outcome. RMD Open. 2023 Apr 17;9(2):e002808. doi:10.1136/rmdopen-2022-002808 PubMed PMID: 37068914; PubMed Central PMCID: PMC10111922.

23. Beswick AD, Wylde V, Gooberman-Hill R, Blom A, Dieppe P. What proportion of patients report long-term pain after total hip or knee replacement for osteoarthritis? A systematic review of prospective studies in unselected patients. BMJ Open. 2012 Jan 1;2(1):e000435. doi:10.1136/bmjopen-2011-000435 PubMed PMID: 22357571.

24. Costello CA, Liu M, Furey A, Rahman P, Randell EW, Zhai G. Association Between Epidemiological Factors and Nonresponders to Total Joint Replacement Surgery in Primary Osteoarthritis Patients. The Journal of Arthroplasty. 2021 May 1;36(5):1502–1510.e5. doi:10.1016/j.arth.2020.11.020

25. Srinivas S, Paquet J, Bailey C, Nataraj A, Stratton A, Johnson M, et al. Effect of spinal decompression on back pain in lumbar spinal stenosis: a Canadian Spine Outcomes Research Network (CSORN) study. Spine J. 2019 Jun;19(6):1001–8. doi:10.1016/j.spinee.2019.01.003 PubMed PMID: 30664950.

26. Lønne G, Fritzell P, Hägg O, Nordvall D, Gerdhem P, Lagerbäck T, et al. Lumbar spinal stenosis: comparison of surgical practice variation and clinical outcome in three national spine registries. Spine J. 2019 Jan;19(1):41–9. doi:10.1016/j.spinee.2018.05.028 PubMed PMID: 29792994.

27. Cooper C, Adachi JD, Bardin T, Berenbaum F, Flamion B, Jonsson H, et al. How to define responders in osteoarthritis. Curr Med Res Opin. 2013 Jun;29(6):719–29. doi:10.1185/03007995.2013.792793 PubMed PMID: 23557069; PubMed Central PMCID: PMC3690437.

28. Costello CA, Hu T, Liu M, Zhang W, Furey A, Fan Z, et al. Metabolomics Signature for Non-Responders to Total Joint Replacement Surgery in Primary Osteoarthritis Patients: The Newfoundland Osteoarthritis Study. Journal of Orthopaedic Research. 2020;38(4):793–802. doi:10.1002/jor.24529

29. Escobar A, Quintana JM, Bilbao A, Aróstegui I, Lafuente I, Vidaurreta I. Responsiveness and clinically important differences for the WOMAC and SF-36 after total knee replacement. Osteoarthritis Cartilage. 2007 Mar;15(3):273–80. doi:10.1016/j.joca.2006.09.001 PubMed PMID: 17052924.

30. Quintana JM, Escobar A, Bilbao A, Arostegui I, Lafuente I, Vidaurreta I. Responsiveness and clinically important differences for the WOMAC and SF-36 after hip joint replacement. Osteoarthritis Cartilage. 2005 Dec;13(12):1076–83. doi:10.1016/j.joca.2005.06.012 PubMed PMID: 16154777.

31. Copay AG, Glassman SD, Subach BR, Berven S, Schuler TC, Carreon LY. Minimum clinically important difference in lumbar spine surgery patients: a choice of methods using the Oswestry Disability Index, Medical Outcomes Study questionnaire Short Form 36, and pain scales. Spine J. 2008;8(6):968–74. doi:10.1016/j.spinee.2007.11.006 PubMed PMID: 18201937.

32. Marsh J, Joshi I, Somerville L, Vasarhelyi E, Lanting B. Health care costs after total knee arthroplasty for satisfied and dissatisfied patients. Can J Surg. 2022 Sep 1;65(5):E562–6. doi:10.1503/cjs.006721 PubMed PMID: 36302132; PubMed Central PMCID: PMC9451500.

33. Escobar A, Gonzalez M, Quintana JM, Vrotsou K, Bilbao A, Herrera-Espiñeira C, et al. Patient acceptable symptom state and OMERACT-OARSI set of responder criteria in joint replacement. Identification of cut-off values. Osteoarthritis Cartilage. 2012 Feb;20(2):87–92. doi:10.1016/j.joca.2011.11.007 PubMed PMID: 22155074.

34. Choi JK, Geller JA, Yoon RS, Wang W, Macaulay W. Comparison of Total Hip and Knee Arthroplasty Cohorts and Short-Term Outcomes From a Single-Center Joint Registry. The Journal of Arthroplasty. 2012 Jun 1;27(6):837–41. doi:10.1016/j.arth.2012.01.016

35. Bourne RB, Chesworth B, Davis A, Mahomed N, Charron K. Comparing Patient Outcomes After THA and TKA: Is There a Difference? Clin Orthop Relat Res. 2010 Feb;468(2):542–6. doi:10.1007/s11999-009-1046-9 PubMed PMID: 19760472; PubMed Central PMCID: PMC2806999.

36. Daher M, Liu J, Baroudi M, Alsoof D, Balmaceno-Criss M, Diebo BG, et al. Patient-reported Physical and Mental Health Outcomes Following Lumbar Spinal Fusion versus Total Hip and Total Knee Replacement. World Neurosurg. 2024 Nov;191:e289–95. doi:10.1016/j.wneu.2024.08.106 PubMed PMID: 39186976.

37. Rampersaud YR, Perruccio AV, Collett E, Sundararajan K, Montoya L, Power JD, et al. Cost-Utility Analysis: Understanding The Economic Impact Of Surgical Non-Response In Orthopaedic HIP, Knee And Spine Surgery For Osteoarthritis. Osteoarthritis and Cartilage. 2023 Mar 1;31:S241–2. doi:10.1016/j.joca.2023.01.240

38. Glassman SD, Copay AG, Berven SH, Polly DW, Subach BR, Carreon LY. Defining substantial clinical benefit following lumbar spine arthrodesis. J Bone Joint Surg Am. 2008 Sep;90(9):1839–47. doi:10.2106/JBJS.G.01095 PubMed PMID: 18762642.

39. Power JD, Perruccio AV, Canizares M, McIntosh G, Abraham E, Attabib N, et al. Determining minimal clinically important difference estimates following surgery for degenerative conditions of the lumbar spine: analysis of the Canadian Spine Outcomes and Research Network (CSORN) registry. Spine J. 2023 Sep;23(9):1323–33. doi:10.1016/j.spinee.2023.05.001 PubMed PMID: 37160168.

40. Berliner JL, Brodke DJ, Chan V, SooHoo NF, Bozic KJ. John Charnley Award: Preoperative Patient-reported Outcome Measures Predict Clinically Meaningful Improvement in Function After THA. Clin Orthop Relat Res. 2016 Feb;474(2):321–9. doi:10.1007/s11999-015-4350-6 PubMed PMID: 26201420; PubMed Central PMCID: PMC4709271.

41. Ontario Ministry of Health. Ontario Case Costing Initiative (OCCI) - Dataset [Internet]. [cited 2025 Jun 17]. Available from: https://data.ontario.ca/en/dataset/ontario-case-costing-initiative-occi

42. Rampersaud YR, Tso P, Walker KR, Lewis SJ, Davey JR, Mahomed NN, et al. Comparative outcomes and cost-utility following surgical treatment of focal lumbar spinal stenosis compared with osteoarthritis of the hip or knee: part 2--estimated lifetime incremental cost-utility ratios. Spine J. 2014 Feb 1;14(2):244–54. doi:10.1016/j.spinee.2013.11.011 PubMed PMID: 24239803.

43. Rampersaud YR, Lewis SJ, Davey JR, Gandhi R, Mahomed NN. Comparative outcomes and cost-utility after surgical treatment of focal lumbar spinal stenosis compared with osteoarthritis of the hip or knee—part 1: long-term change in health-related quality of life. The Spine Journal. 2014 Feb 1;14(2):234–43. doi:10.1016/j.spinee.2013.12.010 PubMed PMID: 24325880.

44. Jentzsch T, Sundararajan K, Rampersaud YR. The clinical course of symptoms during wait time for lumbar spinal stenosis surgery and its effect on postoperative outcome: a retrospective cohort study. Spine J. 2024 Apr;24(4):644–9. doi:10.1016/j.spinee.2023.11.006 PubMed PMID: 38008188.

45. Lebedeva Y, Churchill L, Marsh J, MacDonald SJ, Giffin JR, Bryant D. Wait times, resource use and health-related quality of life across the continuum of care for patients referred for total knee replacement surgery. Canadian Journal of Surgery. 2021 Jun 1;64(3):E253–64. doi:10.1503/cjs.003419 PubMed PMID: 33908239.

46. Blom AW, Donovan RL, Beswick AD, Whitehouse MR, Kunutsor SK. Common elective orthopaedic procedures and their clinical effectiveness: umbrella review of level 1 evidence [Internet]. 2021 Jul 8. doi:10.1136/bmj.n1511

47. te Molder MEM, Dowsey MM, Smolders JMH, van Steenbergen LN, van den Ende CHM, Heesterbeek PJC. Inadequate Classification of Poor Response After Total Knee Arthroplasty: A Comparative Analysis of 15 Definitions Using Data From the Dutch Arthroplasty Register and the Osteoarthritis Initiative Database. The Journal of Arthroplasty. 2024 Oct 1;39(10):2483–9. doi:10.1016/j.arth.2024.05.032

48. Cheng HY, Beswick AD, Bertram W, Siddiqui MA, Gooberman-Hill R, Whitehouse MR, et al. What proportion of people have long-term pain after total hip or knee replacement? An update of a systematic review and meta-analysis. BMJ Open. 2025 May;15(5):e088975. doi:10.1136/bmjopen-2024-088975

49. Turcotte JJ, Brennan JC, Johnson AH, Patton CM. Recovery Trajectories After Lumbar Fusion Stratified by Baseline Patient-Reported Outcomes Measurement Information System Physical Function Disability Levels. Int J Spine Surg. 2025 Apr 24;19(2):207–15. doi:10.14444/8755 PubMed PMID: 40306964; PubMed Central PMCID: PMC12230312.

50. Hatakka J, Laaksonen I, Kostensalo J, Mäkelä KT, Salo H, Pernaa K. 1-year results of lumbar spinal stenosis surgery in Finland: a national FinSpine register study. Acta Orthopaedica. 2025 Feb 14;96:154–60. doi:10.2340/17453674.2025.42849

51. Bech-Azeddine R, Fruensgaard S, Andersen M, Carreon LY. Outcomes of decompression without fusion in patients with lumbar spinal stenosis and substantial back pain [Internet]. 2021 Jan 22. doi:10.3171/2020.8.SPINE20684

52. Sunderland G, Foster M, Dheerendra S, Pillay R. Patient-Reported Outcomes Following Lumbar Decompression Surgery: A Review of 2699 Cases. Global Spine J. 2021 Mar;11(2):172–9. doi:10.1177/2192568219896541 PubMed PMID: 32875849; PubMed Central PMCID: PMC7882820.

53. Gleadhill C, Dooley K, Kamper SJ, Manvell N, Corrigan M, Cashin A, et al. What does high value care for musculoskeletal conditions mean and how do you apply it in practice? A consensus statement from a research network of physiotherapists in New South Wales, Australia [Internet]. 2023 Jun 1. doi:10.1136/bmjopen-2022-071489

54. Mari K, Dégieux P, Mistretta F, Guillemin F, Richette P. Cost utility modeling of early vs late total knee replacement in osteoarthritis patients. Osteoarthritis Cartilage. 2016 Dec;24(12):2069–76. doi:10.1016/j.joca.2016.07.013 PubMed PMID: 27492465.

55. Xie F, Lo NN, Tarride JE, O’Reilly D, Goeree R, Lee HP. Total or partial knee replacement? Cost-utility analysis in patients with knee osteoarthritis based on a 2-year observational study. Eur J Health Econ. 2010 Feb;11(1):27–34. doi:10.1007/s10198-009-0154-5 PubMed PMID: 19430952.

56. Neumann PJ, Kim DD. Cost-effectiveness Thresholds Used by Study Authors, 1990-2021. JAMA. 2023 Apr 18;329(15):1312–4. doi:10.1001/jama.2023.1792

57. Crawford EJ, Ravinsky RA, Coyte PC, Rampersaud YR. Lifetime incremental cost–utility ratios for minimally invasive surgery for degenerative lumbar spondylolisthesis relative to failed medical management compared with total hip and knee arthroplasty for osteoarthritis. Can J Surg. 2021 Jul 23;64(4):E391–402. doi:10.1503/cjs.015719 PubMed PMID: 34296707; PubMed Central PMCID: PMC8410474.

58. Pickens GT, Liang L, Roemer M. HCUP Cost-to-Charge Ratio Methodologies [HCUP Methods Series Report] [Internet]. U.S. Agency for Healthcare Research and Quality; 2021 Dec. (HCUP Methods Series Report). HCUP Methods Series Report nos.: 2021–05. Available from: https://hcup-us.ahrq.gov/reports/methods/MS2021-05-CCR-Methodologies.pdf

59. Dowsey MM, Spelman T, Choong PFM. A Nomogram for Predicting Non-Response to Surgery One Year after Elective Total Hip Replacement. Journal of Clinical Medicine. 2022 Jan;11(6):6. doi:10.3390/jcm11061649

60. Bozic KJ. Improving Value in Healthcare. Clinical Orthopaedics and Related Research®. 2013 Feb;471(2):368. doi:10.1007/s11999-012-2712-x

61. Gandhi R, Tso P, Davis A, Mahomed NN. Outcomes of total joint arthroplasty in academic versus community hospitals. Can J Surg. 2009 Oct;52(5):413–6. PubMed PMID: 19865577; PubMed Central PMCID: PMC2769099.

